# Implications of Monsoon Season & UVB Radiation for COVID-19 in India Manuscript

**DOI:** 10.1101/2020.09.24.20200576

**Authors:** Rahul Kalippurayil Moozhipurath, Lennart Kraft

**Affiliations:** Faculty of Economics and Business, Goethe University Frankfurt

## Abstract

**Background:** India has recorded 66,333 deaths over 36 administrative regions placing India third in the world after the US and Brazil for COVID-19 deaths as of 2 September 2020. Studies indicate that south-west monsoon season plays a role in the dynamics of contagious diseases, which tend to peak post-monsoon season. Recent studies show that vitamin D and its primary source Ultraviolet-B radiation (UVB) may play a protective role in mitigating COVID-19 deaths. However, the combined roles of the monsoon season and UVB in COVID-19 in India are still unclear. In this observational study, we empirically study the respective roles of monsoon season and UVB, whilst further exploring, whether monsoon season negatively impacts the protective role of UVB in COVID-19 deaths in India.

**Methods:** We use a log-linear Mundlak model to a panel dataset of 36 administrative regions in India from 14 March 2020 - 8 August 2020 (n=4005). We use the cumulative COVID-19 deaths as the dependent variable. We isolate the association of monsoon season and UVB as measured by Ultraviolet Index (UVI) from other confounding time-constant and time-varying region-specific factors.

**Findings:** After controlling for various confounding factors, we observe that the monsoon season and a unit increase in UVI are separately associated with 12.8 percentage points and 2.0 percentage points decline in growth rates of COVID-19 deaths in the long run. These associations translate into substantial relative changes. For example, the current monsoon season, that has been going on for two weeks, is associated with a reduction in growth rates of COVID-19 deaths of 59%, whereas a permanent unit increase of UVI is associated with a reduction in growth rates of COVID-19 deaths of 37%. However, the current monsoon season, also reduces the protective role of UVI by 16.3% [0.33 percentage points], plausibly due to lower UVB exposure.

**Interpretation:** We find independent protective roles of both the monsoon season and UVI in mitigating COVID-19 deaths. Furthermore, we find evidence that monsoon season is associated with a significant reduction in the protective role of UVI. The protective role of monsoon season is plausibly due to limited outdoor activities of people. Our study outlines the role of the monsoon season and UVB in COVID-19 in India and supports health-related policy decision making in India.

## 1 Introduction

COVID-19 has caused unparalleled economic and health disruptions in India, the second most populated country with over 1.3 billion people. As of September 2, India has reported 66,333 COVID-19 deaths across 36 administrative regions, placing India third in the world behind the US and Brazil^1^.

Recent observational and clinical studies show that vitamin D deficiency might be linked to incidence^2,3^, severity^4 5^and mortality^6–8^ associated with COVID-19. Recent studies show that vitamin D and its primary source Ultraviolet-B radiation (UVB) may play a protective role in mitigating COVID-19 deaths^9,10^. Studies indicate that south-west monsoon season (monsoon season) plays a role in the dynamics of contagious diseases, which tend to peak post-monsoon season ^11^. The respective roles of the monsoon season and UVB in COVID-19 in India are still unclear. We anticipate a sudden increase in contagious diseases during and post monsoon season, which will stress India’s healthcare system^12,13^. Limited hours of sunlight and dense cloud cover^14^ limit the intensity of UVB radiation, mitigating its protective role^9^ during the monsoon season. Even though limited outdoor activities during the monsoon season may decrease the likelihood of transmission; we anticipate that it may also lead to a lower likelihood of UVB exposure, further mitigating its protective role. To the best of our knowledge, so far, no empirical study has explored the roles of the monsoon season and UVB in COVID-19 in India, specifically studying the association between monsoon season, the subsequent reduced exposure likelihood of UVB radiation and COVID-19 deaths in India.

In this observational study, we empirically describe the roles of the monsoon season and further explore, whether monsoon season result in a reduction in the protective role of UVB in COVID-19 deaths in India. After controlling for various confounding factors, we observe that in the long run the monsoon season and a unit increase in UVI are separately associated with 12.8 percentage points [p<0.05] and 2.0 percentage points [p<0.05] decline in COVID-19 deaths growth rate. On the other hand, in the long run the monsoon season reduces the protective role of UVI by 1.3 percentage points [p<0.05], plausibly due to lower UVB exposure. Consequently, it is expected that although the monsoon season is helping mitigate the transmission of Covid-19, it is doing so at the cost of UVB exposure.

## 2 Impact of Monsoon on Healthcare System, UVB Radiation & COVID-19 Deaths in India

Studies indicate that the monsoon season and post-monsoon season may be associated with the peaks of contagious diseases like influenza, i.e., July-September^11,15,16^. Heavy rainfall linked to monsoon season may create situations favourable for the outbreaks of infectious diseases such as diarrheal disease, cholera, dengue, typhoid as well as respiratory diseases^17^. Furthermore, the temporal overlap between these contagious diseases and COVID-19 may give rise to significant health care challenges^12^. The consequences of possible coinfection with these infectious diseases and SARS-CoV-2 are largely unknown^12,13^. Moreover, we anticipate this sudden increase in contagious diseases during monsoon season to create stress in the healthcare system, further restricting the hospital capacity required for COVID-19 patients^12,13^. Heavy precipitation may also cause disruptions in traffic, limiting the transportation possibilities of COVID-19 patients^13^. On the other hand, the monsoon season also plays a protective role due to restricted mobility^18^ as people are more likely to stay indoors, reducing the possibility of the transmission of the virus. In general, the impact of the monsoon season on COVID-19 in India remains largely unknown.

In addition to the above consequences in the healthcare system, another important consequence of the monsoon season is the higher precipitation and the reduced likelihood of UVB exposure and subsequently lower vitamin D levels^19^. Studies indicate that UV radiation inactivates viruses in fomite transmission^20^. UVB also plays another protective role via its role in vitamin D skin synthesis^21–25^, as dietary intake (natural food, fortified food or supplements) are usually insufficient^26^. Even in a country like India with plenty of sunshine, vitamin D deficiency is common due to reduced skin exposure and specific dietary habits such as vegetarianism^27^. UVB Radiation and the likelihood of skin exposure & skin synthesis vary substantially depending on several factors such as seasons^26^, time^26^, latitude^26^, altitude^26^, active lifestyle^28,29^, dietary habits^30,26^, food fortification^26^, age^26^ and skin colour^26^. Prior studies indicate that most of Indians (Location : 8.4^0^N and 37.6^0^N^31^) belong to skin type V^31^and the time required for recommended vitamin D synthesis for skin type V is 10-15 minutes at solar noon at 11.5^0^N throughout the year^31^. However, this period increases to 10-45 minutes with more duration in winter season at 29^0^N.

Early studies show a protective role of UVB and vitamin D in COVID-19^9,10,2,3,8^. 1,25-dihydroxyvitamin D [1,25 (OH)2D], an active form of vitamin D, plays a critical role in the modulation of innate as well as adaptive immune systems^32,33^, renin-angiotensin system (RAS)^33–35^ as well as in the modulation of the inflammatory response, reducing cytokine storm risk ^32,33^. It also stimulates antimicrobial peptides such as defensins and human cathelicidin^32,33,36,37^ with antiviral properties. Recent studies related to COVID-19 also indicate that vitamin D deficiency might be a risk factor not only for incidence^2,3^, but also for severity^4^ 5and mortality^6–8^ associated with COVID-19.

In light of this emerging evidence regarding vitamin D and COVID-19, we aim to explore the role of monsoon season as well as how it potentially decreases the protective role of UVB in reducing COVID-19 deaths by reducing the likelihood of exposure to UVB radiation. Specifically, we anticipate that the likelihood of UVB radiation exposure undergoes substantial variation during monsoon season primarily due to lower sunshine hours, thick cloud cover and limited outdoor activities. Although monsoon season is likely to reduce the transmission likelihood of SARS-CoV-2 virus due to reduced mobility & outdoor activities^18^, it also reduces exposure likelihood, increasing the potential vitamin D deficiency^9,10^ among the population. Lower vitamin D levels due to reduced exposure to UVB radiation during monsoon season may lead to lower immunity leading to increased mortality rate^9,26^. Therefore, we anticipate that even though UVB plays a protective role in COVID-19 in India, monsoon season reduces this protective role by reducing the likelihood of exposure.

Studies show that other weather factors such as humidity^38^, temperature^39^and precipitation^39,40^ play a role in viral transmission (e.g., influenza). Recent studies also indicate that temperature and humidity may play a role in COVID-19 transmission^40,41^. Therefore, we control for most of these confounding weather factors to explore the role of monsoon season as well as how it potentially decreases the protective role of UVB in reducing COVID-19 deaths.

## 3 Methods

### 3.1 Description of Data

In order to identify the relation of monsoon season, UVI and their interaction with COVID-19 deaths, we constructed the dataset outlined in Table 1. We collected COVID-19 data across 36 administrative regions (28 states and 8 union territories) of India covering 148 days from 14 March 2020 until 8 August 2020. 34 of these administrative regions reported more than 20 COVID-19 infections on 8 August 2020. We focus on those 34 administrative regions to ensure that the results are not biased by those administrative regions that are at a very early stage of the COVID-19 outbreak, which would limit data points concerning COVID-19 deaths. Furthermore, we drop the first 20 daily observations of every administrative region after the reporting of the first COVID-19 infection in respective regions. Thus, we ensure that these observations at the very early stage of the outbreak do not bias the results.

**Table 1:** Summary of dataset.

|  |  |
| --- | --- |
| Number of administrative regions of India in our data set | 36 |
| ... > 0 cumulated number of COVID-19 deaths before 8 August 2020 | 34 |
| ... > 20 cumulated number of COVID-19 infections before 8 August 2020 | 34 |
| Covered time-period | 14 March 2020 - 8 August 2020 (148 days) |
| Granularity of data | Daily |
| COVID-19 data source | https://www.covid19india.org/ |
| Latitude and longitude data source for each state that is used to match weather data | Geocoder (Python) |
| Weather data source | https://darksky.net/ |
| Monsoon season data source | https://indianexpress.com/article/india/from-june-2020-revised-monsoon-calendar-for-india-6364258/<br>https://mausam.imd.gov.in/imd_latest/contents/monsoon.php |

The corresponding data consist of the cumulative daily number of COVID-19 deaths at an administrative region level. They also consist of the time of arrival of monsoon season for each administrative region, the daily ultraviolet index (UVI), an indicator of daily UVB radiation, as well as a set of daily weather parameters. We use these weather parameters as control variables, and these parameters include cloud index, ozone level, visibility level, humidity level, minimum and maximum temperature. We source COVID-19 data from COVID19India.org and the weather data from darksky.net based on the latitude and longitude information of countries that are provided by Geocoder, a geocoding library in Python.

We present descriptive statistics of the dataset in Table 2. As of 8 August 2020, the cumulative COVID-19 deaths of these 34 administrative regions were on average 1,300, and the growth rate of COVID-19 deaths across all administrative regions on 8 August was on average 3.5% as compared to the average growth rate of COVID-19 deaths across all administrative regions and time which was 5.4%. On average, the first reported COVID-19 infection in each administrative region happened 137 days before 8 August 2020. The monsoon season had arrived in 33 of 34 administrative regions as of 8 August 2020. UVI is on average 8.5 as of this date.

**Table 2:** Descriptive statistics of data set.

| Variable | Number of State | Number of Observations | Mean | Std. Dev. | Min | Max |
| --- | --- | --- | --- | --- | --- | --- |
| Cumulated COVID-19 deaths on 8 August 2020 | 34 | 34 | 1300 | 3100 | 1 | 17,400 |
| Growth rate of cumulative COVID-19 deaths on 8 August | 34 | 34 | 0.035 | 0.037 | 0 | 0.2 |
| Daily growth rate of cumulative COVID-19 deaths | 34 | 3,227 | 0.054 | 0.15 | -0.5 | 3 |
| Time-passed by from first reported infection until 8 August 2020 | 34 | 34 | 137 | 19 | 76 | 148 |
| Monsoon arrived by 8 August 2020 | 34 | 34 | 0.97 | 0.17 | 0 | 1 |
| Daily Ultraviolet index (UVI) | 34 | 4,005 | 8.5 | 2.7 | 4 | 15 |
| Daily cloud index | 34 | 4,005 | 0.60 | 0.32 | 0 | 1 |
| Daily ozone level | 34 | 4,005 | 278 | 14 | 254 | 343 |
| Daily visibility level | 34 | 4,005 | 15 | 1.7 | 1.0 | 16 |
| Daily humidity level | 34 | 4,005 | 0.71 | 0.22 | 0.09 | 1 |
| Minimum temperature per day within a country | 34 | 4,005 | 22 | 6.8 | -14 | 32 |
| Maximum temperature per day within a country | 34 | 4,005 | 31 | 7.6 | -2.5 | 46 |

### 3.2 Description of Methodology

We use a Mundlak error correction model to estimate the association of monsoon season, UVI and their interaction with COVID-19 deaths. We include 56 days moving average of the monsoon season dummy variables, which indicate whether the monsoon season has been active in a region on a specific day. We also use 56 days moving averages of the UVI Index as well as of the interaction of monsoon season dummy variables and UVI Index for a region on a specific day.

Our model aims to isolate all weather parameters from region-specific time-constant factors. The Mundlak model combines the robustness of a fixed-effects model and the efficiency of a random-effects model. Instead of demeaning the structural model to isolate the weather parameters analytically from such region-specific time-constant factors, it models those region-specific time-constant factors through the available weather parameters. We describe our methodology and how to interpret the estimated associations in more detail in *Supplementary Material* section 1 and section 2.

## 4 Results

Table 3 outlines our main results. We find substantial and significant associations of monsoon season, UVI and their interaction with COVID-19 deaths. Monsoon season is associated with a drop of 12.8 percentage points [p<0.05] in daily growth rates of COVID-19 deaths in the long run. A permanent unit increase of UVI is associated with a decline of 2 percentage points [p<0.05] in daily growth rates of COVID-19 deaths. However, monsoon season mitigates the association of UVI by 1.3 percentage points [p<0.05] and thus alleviates the protective role of UVI.

**Table 3:**
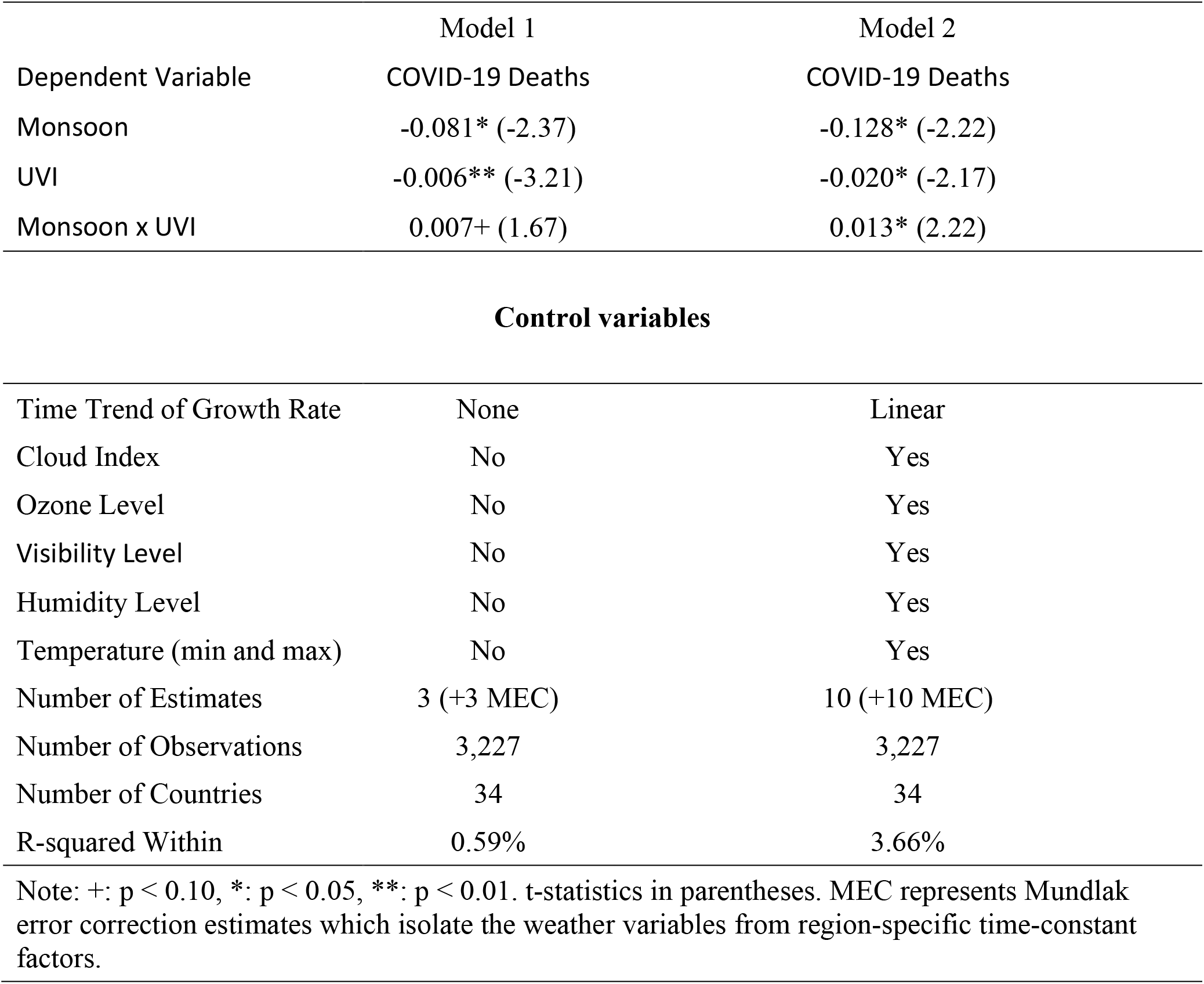
Effect of Monsoon Season, UVI and their interaction on cumulative COVID-19 deaths.

|  | Model 1 | Model 2 |
| --- | --- | --- |
| Dependent Variable | COVID-19 Deaths | COVID-19 Deaths |
| Monsoon | -0.081* (-2.37) | -0.128* (-2.22) |
| UVI | -0.006** (-3.21) | -0.020* (-2.17) |
| Monsoon x UVI | 0.007+ (1.67) | 0.013* (2.22) |

| Control variables |  |  |
| --- | --- | --- |
| Time Trend of Growth Rate | None | Linear |
| Cloud Index | No | Yes |
| Ozone Level | No | Yes |
| Visibility Level | No | Yes |
| Humidity Level | No | Yes |
| Temperature (min and max) | No | Yes |
| Number of Estimates | 3 (+3 MEC) | 10 (+10 MEC) |
| Number of Observations | 3,227 | 3,227 |
| Number of Countries | 34 | 34 |
| R-squared Within | 0.59% | 3.66% |

Note: +: $p < 0.10$ , \*: $p < 0.05$ , \*\*: $p < 0.01$ . t-statistics in parentheses. MEC represents Mundlak error correction estimates which isolate the weather variables from region-specific time-constant factors.

These associations of monsoon season with COVID-18 deaths translate into substantial relative changes in growth rates of COVID-19 deaths. Within two weeks of the arrival, monsoon season is associated with a reduction of COVID-19 growth rates of 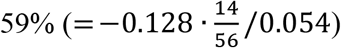 relative to the average daily growth rates of COVID-19 deaths. A permanent unit increase of UVI is associated with a decline in COVID-19 growth rates of 37% (= −0.02/0.054) as relative to the average daily growth rates of COVID-19 deaths. However, after two weeks monsoon season reduces the protective role of UVI by 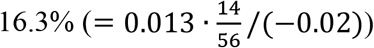. After eight weeks monsoon reduces the protective role of UVI by 65% (= 0.013/(−0.02)). The robustness checks in Table S1 (*Supplementary Material*) outline that our estimations are consistent when using more flexible time trends as well as to the impact of governmental measures on the behaviour of individuals.

## 5 Discussion

Our empirical results outline that monsoon season and UVB are independently associated with a decline in the daily COVID-19 death growth rate in India, thereby indicating their respective protective roles. However, monsoon season also significantly decreases the protective role of UVB radiation. Specifically, we find that monsoon season and a unit increase in UVI are separately associated with -12.8 percentage points, and -2.0 percentage points decline in COVID-19 daily deaths growth rate in India in the long run. In the long run, Indian monsoon season further reduces the protective role of UVI by 1.3 percentage points, potentially due to lower UVB exposure as a result of decreased outdoor activities. We find these results to be consistent across different model specifications.

We control for all time-constant and region-specific factors and different time-varying confounding factors, especially weather^9,10^. We may not be able to exclude other time-varying factors, which might bias our results^9,10^. Further, we may not be able to exclude other protective roles of UVB – for example via other mediators –nitric oxide^9,10^, cis-urocanic acid^21,42,43^ or via inactivation of viruses in fomite transmission^20^. Sensible sunlight exposure^9^ is crucial as disproportionate solar UV exposure leads to health hazards such as aging^44^, wrinkles^44^, sunburn^26^ and DNA damage^44^. Specifically, disproportionate UVB exposure is associated with basal cell and squamous cell carcinoma^45^.

We also acknowledge that the results of our study cannot serve as health guidance for India. However, we hope our results prompt further clinical research in India specifically to establish the role of sensible sunlight exposure or vitamin D in mitigating COVID-19 deaths during monsoon season. Establishing the effectiveness of sensible solar UVB exposure or vitamin D supplementation could substantially advance the control of COVID-19 pandemic at scale in India. The results of these clinical studies can further guide policy decision making in India, especially during monsoon season. This type of policy intervention would be desirable for India not only due to its lower risk and costs, but also due to its scalability across India’s 1.3 billion people whose economic means vary significantly.

## Data Availability

The data used in the study are from publicly available sources. Data regarding COVID-19 are obtained on 9 August 2020 from https://www.covid19india.org/. We will make specific data set used in this study available for any future research. Interested researchers can contact one of the authors via email to get access to the data.

## Acknowledgements

We would like to thank Bernd Skiera for providing support to this paper. We would like to acknowledge Sharath Mandya Krishna - for providing inputs and assisting with data collection, data transformation and data engineering. We thank Matthew Little for his inputs and his assistance in review. We would also like to acknowledge Magdalena Ceklarz for her valuable contributions to our paper and the discussions about COVID-19 at different points in time.

## 8 Author Contributions

RKM conceptualized the research idea, conducted literature research, designed theoretical framework and collected COVID-19 and weather data. LK collected data regarding monsoon season. LK designed empirical methods and analyzed the data. RKM and LK interpreted the results and wrote the article.

## 10 Additional Information

Correspondence and requests for materials should be addressed to Rahul Kalippurayil Moozhipurath.

## 11 Data Sharing

The data used in the study are from publicly available sources. Data regarding COVID-19 are obtained on 9 August 2020 from https://www.covid19india.org/. Data regarding weather is obtained from *Dark Sky* on the 9 August 2020 and can be accessed at https://darksky.net/. Latitude and longitude information is obtained via Geocoder (Python), whereas monsoon season data is obtained from https://indianexpress.com/article/india/from-june-2020-revised-monsoon-calendar-for-india-6364258/ as well as from https://mausam.imd.gov.in/imd_latest/contents/monsoon.php on the 9 August 2020. We will make specific data set used in this study available for any future research. Interested researchers can contact one of the authors via email to get access to the data.

